# Traumatic Brain Injury and Risk of Cardiometabolic Multimorbidity: a Prospective Cohort Study

**DOI:** 10.64898/2026.05.07.26352704

**Authors:** Shuai Li, Xiangbin Liu, Xiaoyue Chen, Yuerong Liu, Li Lin, Siyu Liu, Chuanchang Li, Yongping Bai, Wei Xie, Xunjie Cheng

## Abstract

**Background:** Recent studies have established an association between traumatic brain injury (TBI) and cardiometabolic diseases (CMDs). However, the influence of TBI on the sequential progression from a healthy state to CMD, subsequent to cardiometabolic multimorbidity (CMM), and ultimately to mortality remains unclear.

**Methods:** A total of 366,616 participants free of CMD at baseline were derived from the UK Biobank (UKB). CMM was defined as the co-occurrence of ≥2 CMD, including diabetes mellitus (DM), ischemic heart disease (IHD), and stroke. Cox proportional hazards models and multi-state models were utilized to evaluate the association of TBI with disease transitions from a healthy state to CMM and subsequent mortality.

**Results:** During a median follow-up of 16.91 years, 54,224 participants developed at least one CMD, among whom 7,562 progressed to CMM. Furthermore, 32,785 cases of mortality were documented. In multi-state models, the hazard ratios (HRs) with corresponding 95% confidence intervals (CIs) for transitions from a healthy state to IHD, DM, stroke, and mortality were 1.91(95% CI: 1.77–2.05), 1.89 (95% CI: 1.71–2.09), and 4.73 (95% CI: 4.39–5.09), respectively. For sequential transitions from IHD, DM, and stroke to CMM, the HRs (95% CIs) were 2.67 (95% CI: 2.34–3.04), 3.29 (95% CI: 2.78–3.89), and 1.41 (95% CI: 1.15–1.72), respectively. Additionally, in Cox proportional hazards models, the HRs (95% CIs) for incident CMM and mortality among individuals with TBI were 3.98 (95% CI: 3.63–4.36) and 2.57 (95% CI: 2.44–2.71), respectively.

**Conclusion:** This study found that TBI was associated with increased risk of progression from a healthy state to CMD, and subsequently to CMM and mortality, highlighting the importance of comprehensive management of TBI in cardiometabolic health.

**What is Known; What the Study Adds:** *What is Known:* Traumatic brain injury (TBI) is associated with an elevated risk of developing multiple cardiometabolic diseases (CMDs).

*What the Study Adds:* This study performed a systematic analysis of the relationships between TBI and multiple CMDs, providing valuable clinical references for the prevention and management of the onset and progression of cardiometabolic diseases and cardiometabolic multimorbidity (CMM) among patients with TBI.

## Background

Cardiometabolic multimorbidity (CMM) is defined as the co-occurrence of ≥2 cardiometabolic diseases (CMDs) in the same individual. CMDs, including diabetes mellitus (DM), ischemic heart disease (IHD), and stroke, represent one of the most consistent manifestations of multimorbidity globally, coinciding with reduced quality of life.^1, 2^ CMM development and progression are linked to multiple factors, including genetics, lifestyle, and comorbid chronic conditions.^3^ Two independent cohort studies^4, 5^ have demonstrated that patients with CMD face significantly elevated risk of mortality or other adverse outcomes compared to those without CMD; Of note, individuals with CMM exhibit even higher risk of adverse outcomes than those with single CMD.^4, 5^ Therefore, identifying modifiable and non-modifiable factors is crucial for preventing CMD onset and its progression to CMM or even mortality.

TBI is a global health problem that causes long-term disability and even mortality in millions of people each year, mainly due to road traffic accidents, falls and violence.^6–9^ TBI-related sequelae include a spectrum of neuropsychological deficits (e.g., cognitive impairment, mood disorders) and increased risk of neurodegenerative diseases, which in turn alter patients’ behavioral and lifestyle patterns.^10,11^ Additionally, TBI has been shown to interact with multisystem chronic diseases, contributing to chronic endocrine dysfunction.^12^ Furthermore, observational evidence indicates that TBI and chronic disease-related metabolic dysfunction synergistically exacerbate neurological outcomes in TBI patients, thereby impairing their overall prognosis.^13^ Collectively, these findings suggest that TBI may represent a key factor associated with CMM, warranting further investigation.

Although the association between TBI and single CMD has been well documented in numerous studies, accumulating evidence indicates that TBI independently elevates the risk of hypertension and stroke.^14, 15^ For instance, a cohort study of US veterans reported that individuals with a TBI history had a higher risk of cardiovascular disease (CVD) than those without TBI: mild TBI was associated with a 62% elevated risk, while moderate-to-severe TBI conferred a 163% increased risk.^16^Potential mechanisms underlying this elevated risk include neuroinflammation and vascular dysfunction; specifically, blood-brain barrier disruption is an established early event following TBI, which may induce chronic neurodegenerative changes and subsequently impair cardiovascular health.^17^ However, the impact of TBI on disease progression trajectories across the CMM spectrum remains understudied. Specifically, the potential differential effects of TBI on individual CMDs at distinct stages of the transition from health to CMM, as well as the subsequent risk of mortality at each stage, remain unelucidated.

Based on about 0.5 million participants from the UK Biobank (UKB), multi-state models were used to investigate the potential differential associations of TBI with the sequential transitions from a healthy state to CMD, subsequent progression to CMM, and eventual mortality. This study aimed to emphasize the importance of comprehensive management of TBI in cardiometabolic health.

## Study Design and Methods

### Study Population

UKB is a large prospective longitudinal biomedical cohort study of 502,505 UK adults aged 37–73 years, recruited across 22 assessment centers between 2006 and 2010.^18^ Data on sociodemographic characteristics, diet, lifestyle, reproductive factors, and environmental exposures were collected, with anthropometric measurements acquired via standardized protocols. From baseline recruitment onward, participants were linked to electronic health records (EHRs) for morbidity and mortality surveillance, using hospitalization and national mortality registries. Detailed study protocols have been described elsewhere.^19, 20^ For the present longitudinal analysis, participants were excluded if they met any of the following criteria: (1) Study withdrawal; (2) Pre-existing CMD or CMM at baseline; (3) Missing TBI data at baseline; (4) Incomplete covariate data. Ultimately, a total of 135,889 participants were excluded, leaving 366,616 participants were included in the final analysis. Each participant provided a written informed consent prior to participation. This study received approved from the Ethics Committee of North West Multi-Centre Research.

### Traumatic brain injury and covariates

TBI was ascertained using multiple data sources from the UKB between 2000 and 2026, including primary care records, hospital admission data, and self-reported medical conditions. Participants were classified as having prevalent TBI at baseline if their TBI diagnosis date preceded study enrollment. Follow-up was conducted from study entry until 2026. Covariates encompassed three categories: (1) demographic characteristics (age, sex, ethnicity); (2) lifestyle factors (smoking status, alcohol consumption, physical activity levels, dietary patterns); and (3) clinical, biochemical, and medication-related variables (body mass index [BMI], systolic blood pressure [SBP], diastolic blood pressure [DBP], pulse rate, insomnia, total cholesterol levels, low-density lipoprotein [LDL] cholesterol levels, and history of using lipid-lowering agents, antihypertensive medications, or aspirin).Detailed information regarding the covariates is provided in additional file 1.

### Outcomes

Disease outcomes, mortality, and time-to-diagnosis were ascertained using the same data sources and methods as TBI status. Specifically, DM was defined by International Classification of Diseases, 10th Revision (ICD-10) codes E11–E14, IHD by codes I20–I25, and stroke by codes I60–I64 and I69. CMD was defined as the presence of any one of these three conditions, while CMM was defined as the co-occurrence of ≥2 of the aforementioned CMDs.^21^ For timeline ascertainment: the date of diagnosis of a first incident CMD was recorded as the time of incident CMD onset; the date of diagnosis of a second CMD was defined as the time of new CMM onset. Follow-up was conducted from study enrollment until the occurrence of the primary outcome, loss to follow-up, death, or March 1, 2026, whichever came first.

### Statistical analysis

Baseline characteristics were stratified by TBI status: continuous variables were presented as mean ± standard deviation (SD), and categorical variables as frequencies (percentages). Kaplan-Meier analysis was used to calculate the cumulative incidence of the first event (isolated CMD, CMM, and mortality), from a healthy state, stratified by TBI status. Cox proportional hazards models were employed to estimate hazard ratios (HRs) and 95% confidence intervals (CIs) for the associations between TBI and isolated CMD, CMM, and mortality, respectively. Model 1 was adjusted for demographic factors (age, gender and race); Model 2 was adjusted for lifestyle factors (smoking status, alcohol consumption, physical activity, and dietary patterns) based on Model 1; Model 3 (fully adjusted model) further incorporated anthropometric, clinical, and biochemical variables (BMI, SBP, DBP, pulse rate, insomnia, total cholesterol levels, and LDL cholesterol levels) in addition to Model 2 adjustments.

Multi-state models were applied to further assess the role of TBI in sequential transitions: from health to CMD, from CMD to CMM, and from CMM to mortality. Given the mutual independence of the three CMD components (IHD, DM, and stroke), 11 distinct transition pathways were generated. Throughout these analyses, CMM was consistently defined as the co-occurrence of ≥2 of the aforementioned CMDs. Subgroup analyses were conducted across all transition pathways, stratified by age (e.g., <65 vs. ≥65 years), sex (male vs. female), and BMI categories (normal vs. obese), to examine effect modification by these factors. Furthermore, to quantify statistical heterogeneity in TBI risk estimates and mitigate potential reverse causality bias, we performed five sensitivity analyses: (1) Exclusion of participants who experienced outcome events within 2 years of baseline recruitment; (2) Exclusion of participants with self-reported disease diagnoses (retaining only registry-confirmed events); (3) Exclusion of cases with multiple outcome events recorded on the same date; (4) Additional adjustment for medication use (antihypertensive agents, lipid-lowering agents, and aspirin); (5) Inclusion of participants with baseline isolated CMD in the CMD-to-CMM and CMD-to-mortality transition models, and participants with baseline CMM in the CMM-to-mortality transition models.

All analyses were performed using R software version 4.2.1 (R Foundation for Statistical Computing, Vienna, Austria). A two-tailed P-value < 0.05 was deemed statistically significant.

## Results

### Descriptive analysis

Baseline characteristics of participants by TBI status are presented in **Table 1**. Of the 366,616 participants, 361,386 were TBI free at baseline and 5,230 were diagnosed with TBI at baseline. Regarding baseline characteristics, participants with TBI were more likely to be male, current or former smokers, and users of antihypertensive agents, lipid-lowering agents, or aspirin; conversely, they were less likely to have insomnia. Furthermore, TBI participants exhibited lower physical activity and dietary pattern scores (all P < 0.01).

**Table 1.**
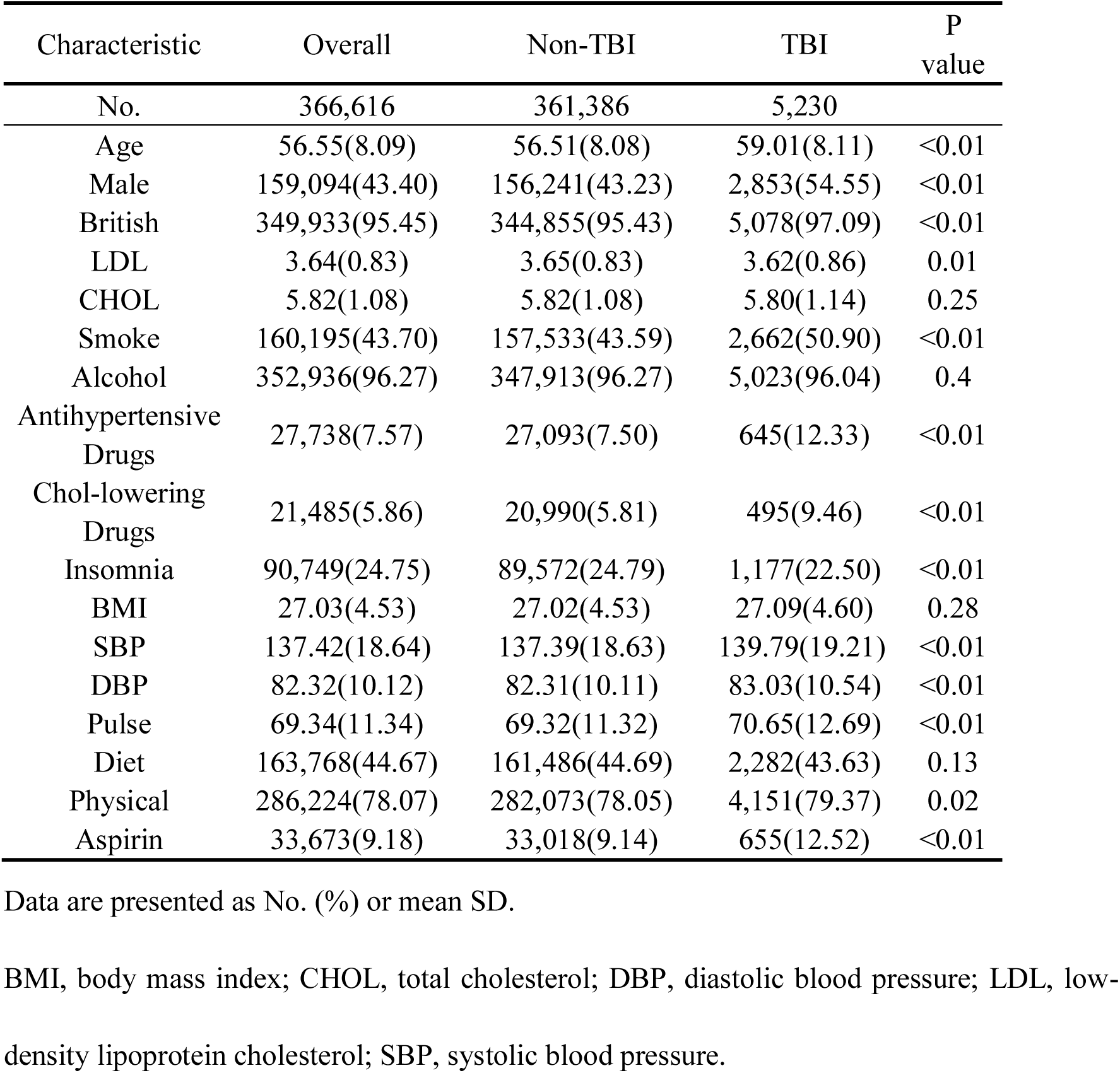
Baseline Characteristics of the Study Population.

| Characteristic | Overall | Non-TBI | TBI | P value |
| --- | --- | --- | --- | --- |
| No. | 366,616 | 361,386 | 5,230 |  |
| Age | 56.55(8.09) | 56.51(8.08) | 59.01(8.11) | <0.01 |
| Male | 159,094(43.40) | 156,241(43.23) | 2,853(54.55) | <0.01 |
| British | 349,933(95.45) | 344,855(95.43) | 5,078(97.09) | <0.01 |
| LDL | 3.64(0.83) | 3.65(0.83) | 3.62(0.86) | 0.01 |
| CHOL | 5.82(1.08) | 5.82(1.08) | 5.80(1.14) | 0.25 |
| Smoke | 160,195(43.70) | 157,533(43.59) | 2,662(50.90) | <0.01 |
| Alcohol | 352,936(96.27) | 347,913(96.27) | 5,023(96.04) | 0.4 |
| Antihypertensive<br>Drugs | 27,738(7.57) | 27,093(7.50) | 645(12.33) | <0.01 |
| Chol-lowering<br>Drugs | 21,485(5.86) | 20,990(5.81) | 495(9.46) | <0.01 |
| Insomnia | 90,749(24.75) | 89,572(24.79) | 1,177(22.50) | <0.01 |
| BMI | 27.03(4.53) | 27.02(4.53) | 27.09(4.60) | 0.28 |
| SBP | 137.42(18.64) | 137.39(18.63) | 139.79(19.21) | <0.01 |
| DBP | 82.32(10.12) | 82.31(10.11) | 83.03(10.54) | <0.01 |
| Pulse | 69.34(11.34) | 69.32(11.32) | 70.65(12.69) | <0.01 |
| Diet | 163,768(44.67) | 161,486(44.69) | 2,282(43.63) | 0.13 |
| Physical | 286,224(78.07) | 282,073(78.05) | 4,151(79.37) | 0.02 |
| Aspirin | 33,673(9.18) | 33,018(9.14) | 655(12.52) | <0.01 |

Data are presented as No. (%) or mean SD.
BMI, body mass index; CHOL, total cholesterol; DBP, diastolic blood pressure; LDL, low-
density lipoprotein cholesterol; SBP, systolic blood pressure.

### Associations between TBI and disease outcomes plus mortality

Regarding clinical outcomes, 54,224 participants were diagnosed with CMD and 7,562 subsequently developed CMM during a median follow-up of 16.91 years. The incidence rates of CMD and CMM were 10.06 and 1.20 per 1,000 person-years, respectively. Kaplan-Meier curves illustrating the cumulative incidence of CMD, CMM, and mortality from survival analyses are presented in **Figure 1**. Cox proportional hazards model results show that fully adjusted HRs values for participants with TBI compared with participants without TBI are: IHD was 1.88 (95% CI, 1.76–2.01), stroke was 4.71 (95% CI, 4.43–5.02), DM was 1.81 (95% CI, 1.66–1.98), CMM was 3.98 (95% CI, 3.63–4.36), and mortality was 2.57 (95% CI, 2.44–2.71)**(Table 2)**

**Figure 1.**
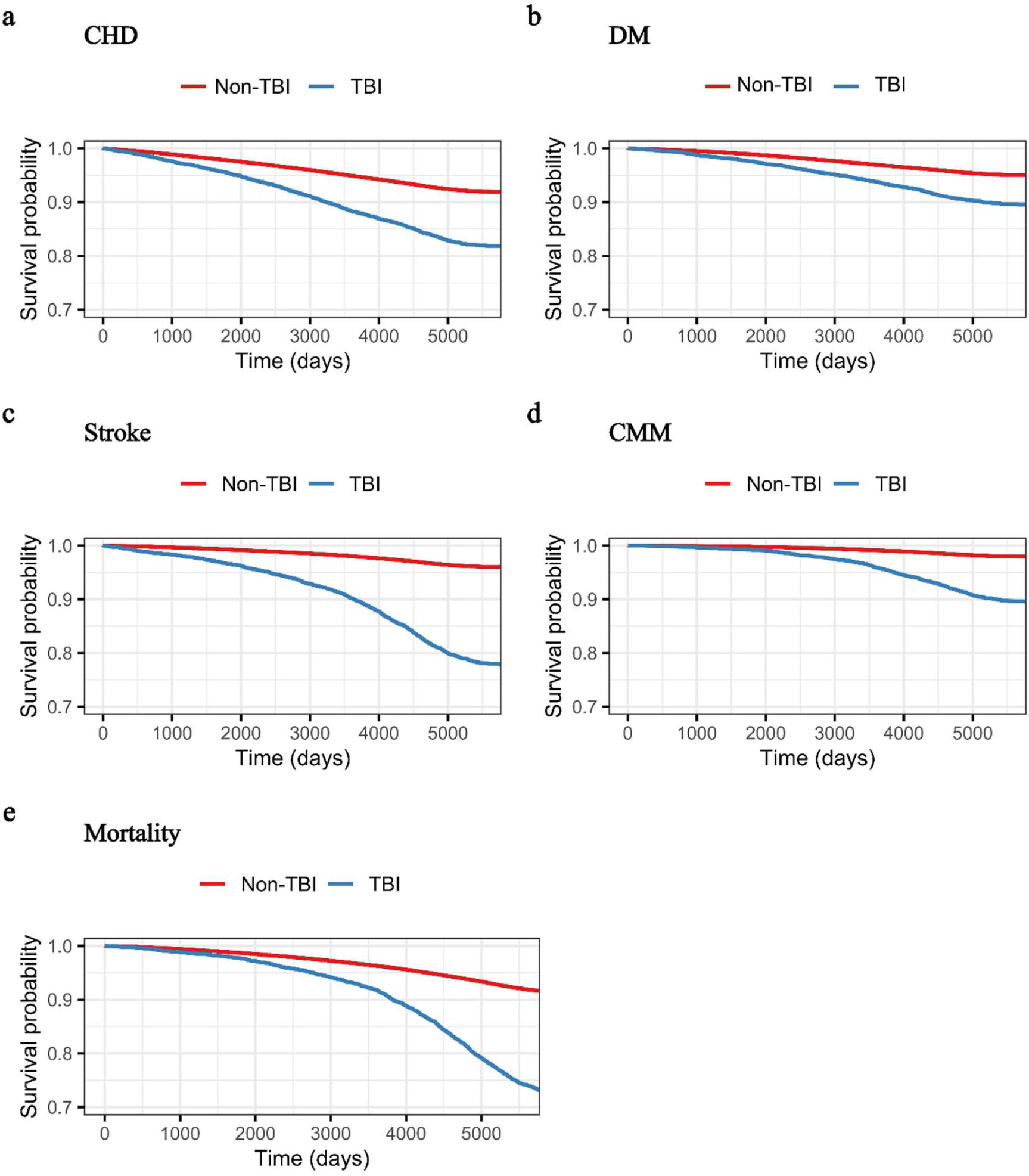
Kaplan-Meier survival curve of people with TBI and non-TBI in the baseline in different outcomes. Multivariate adjusted model hazard ratios for patients with and without TBI who developed IHD (a), DM (b), Stroke (c), CMM (d) and Mortality (e). The solid lines represent survival rates in patients without TBI and the dotted lines represent the survival rate of patients with TBI.

**Table 2.** Cox Analysis Between TBI and four cardiac metabolic outcomes and mortality.

| Variable | Model 1<sup>a</sup> |  | Model 2<sup>b</sup> |  | Model 3<sup>c</sup> |  |
| --- | --- | --- | --- | --- | --- | --- |
|  | HR (95%CI) | P value | HR (95%CI) | P value | HR (95%CI) | P value |
| IHD (n=29,395) |  |  |  |  |  |  |
| NO | Ref | ... | Ref | ... | Ref | ... |
| YES | 1.89 (1.77, 2.02) | <0.01 | 1.86 (1.74, 1.99) | <0.01 | 1.88 (1.76, 2.01) | <0.01 |
| Stroke (n=14,998) |  |  |  |  |  |  |
| NO | Ref | ... | Ref | ... | Ref | ... |
| YES | 4.77 (4.48, 5.08) | <0.01 | 4.73 (4.44, 5.03) | <0.01 | 4.71 (4.43, 5.02) | <0.01 |
| DM (n=17,947) |  |  |  |  |  |  |
| NO | Ref | ... | Ref | ... | Ref | ... |
| YES | 1.92 (1.75, 2.09) | <0.01 | 1.88 (1.72, 2.06) | <0.01 | 1.81 (1.66, 1.98) | <0.01 |
| CMM (n=7,562) |  |  |  |  |  |  |
| NO | Ref | ... | Ref | ... | Ref | ... |
| YES | 4.14 (3.77, 4.54) | <0.01 | 4.06 (3.70, 4.45) | <0.01 | 3.98 (3.63, 4.36) | <0.01 |
| mortality (n=32,785) |  |  |  |  |  |  |
| NO | Ref | ... | Ref | ... | Ref | ... |
| YES | 2.68 (2.54, 2.83) | <0.01 | 2.64 (2.50, 2.78) | <0.01 | 2.57 (2.44, 2.71) | <0.01 |

a Adjusted for age, gender and race
b Adjusted for model 1 plus smoke, alcohol, sport and diet
c Adjusted for model 2 plus cholesterol levels, low-density lipoprotein levels, insomnia, BMI, SBP, DBP and Pulse

### TBI and Multimorbidity: multi-state analysis

Multi-state model analyses demonstrated that TBI exhibited statistically significant associations across all sequential transition stages from a healthy state to CMM and mortality. Specifically, the HRs with corresponding 95% CIs for transitions from a healthy state to IHD, DM, and stroke were 1.91(95% CI: 1.77–2.05), 1.89 (95% CI: 1.71–2.09), and 4.73 (95% CI: 4.39–5.09), respectively. For sequential transitions from IHD, DM, and stroke to CMM, the respective HRs (95% CIs) were 2.67 (95% CI: 2.34–3.04), 3.29 (95% CI: 2.78–3.89), and 1.41 (95% CI: 1.15–1.72). Furthermore, for transitions from a healthy state, IHD, DM, stroke, and CMM to all-cause mortality, the HRs (95% CIs) were 2.21 (95% CI: 2.04–2.39), 1.67 (95% CI: 1.44–1.93), 1.92 (95% CI: 1.54–2.41), 1.43 (95% CI: 1.27–1.61), and 1.68 (95% CI: 1.14–1.92), respectively (Figure 2).

**Figure 2.**
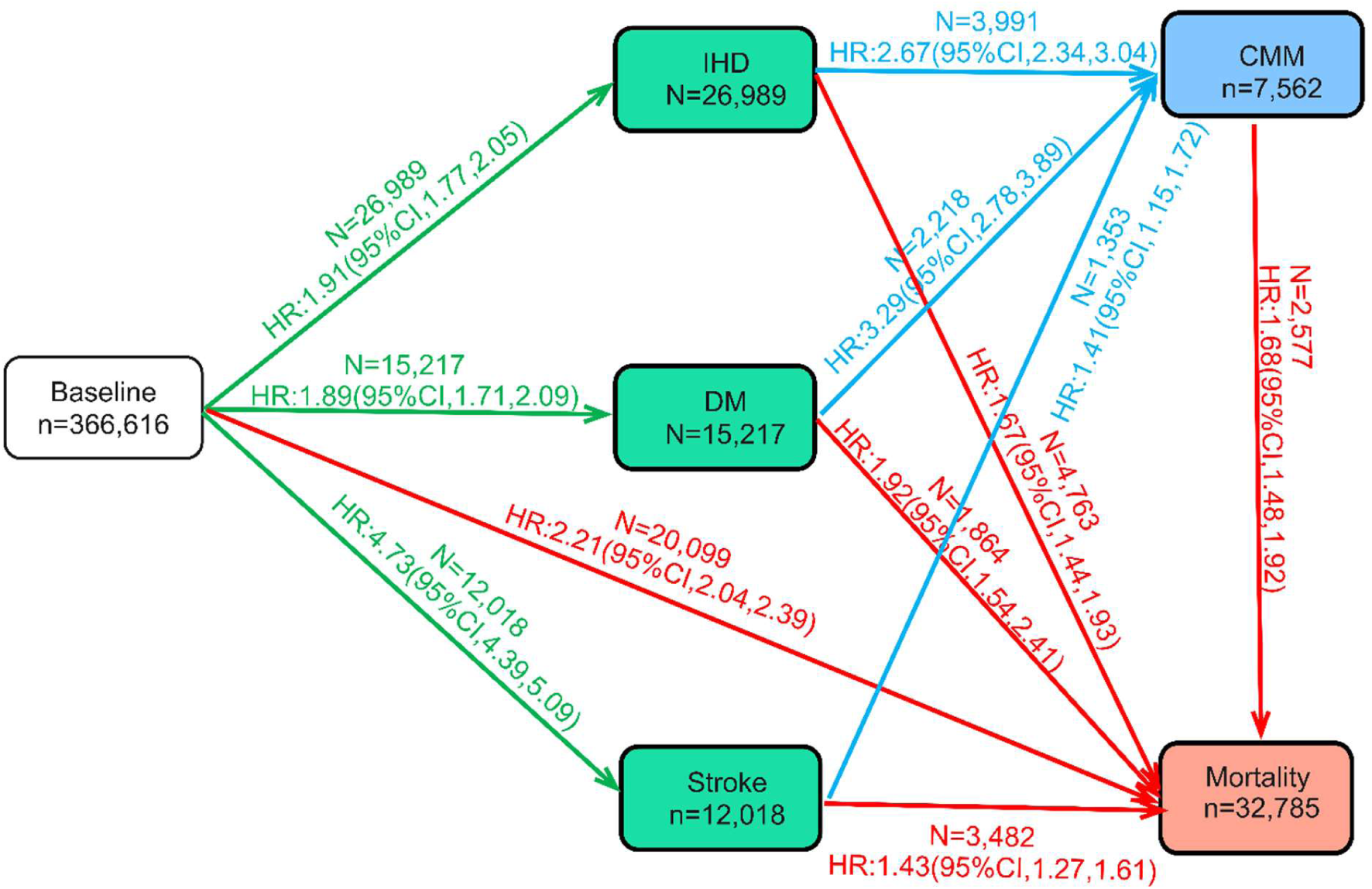
Numbers (percentages) of participants from baseline to one of IHD, DM, and Stroke, Cardiometabolic multimorbidity (CMM), then to Mortality. Cardiometabolic diseases include ischemic heart disease (IHD), Stroke, and diabetes mellitus (DM). CMM is defined as the occurrence of at least two of the above-mentioned diseases. The HRs were reported by TBI vs non-TBI at baseline. Models were adjusted for age at baseline, gender, race, LDL, CHOL, smoke, alcohol, antihypertensive drugs, Chol-lowering drugs, insomnia, BMI, SBP, DBP, pulse, diet, physical. HR=hazard ratio; CI, confidence interval.

### Sensitivity and subgroup analyses

In the sensitivity analyses (Figure 3), results from all supplementary sensitivity tests remained consistent with the overall trends of the primary multi-state model analyses, confirming the statistical robustness of the core findings in this study. Only a few statistically significant interactions were found in stratified analyses, most of which appeared clinically insignificant. Intriguingly, sex-stratified analyses revealed that TBI exerted a stronger impact on the transition from a healthy state to CMD in female participants than in male participants. Conversely, BMI-stratified analyses showed that the strength of the association between TBI and CMD was weaker in the BMI ≥30 kg/m² group compared to the BMI <30 kg/m² group. A similar pattern was observed in age-stratified analyses, with one exception: TBI was associated with a higher risk of transition from stroke to all-cause mortality in individuals aged ≥65 years than in those aged <65 years (Figure 4).

**Figure 3.**
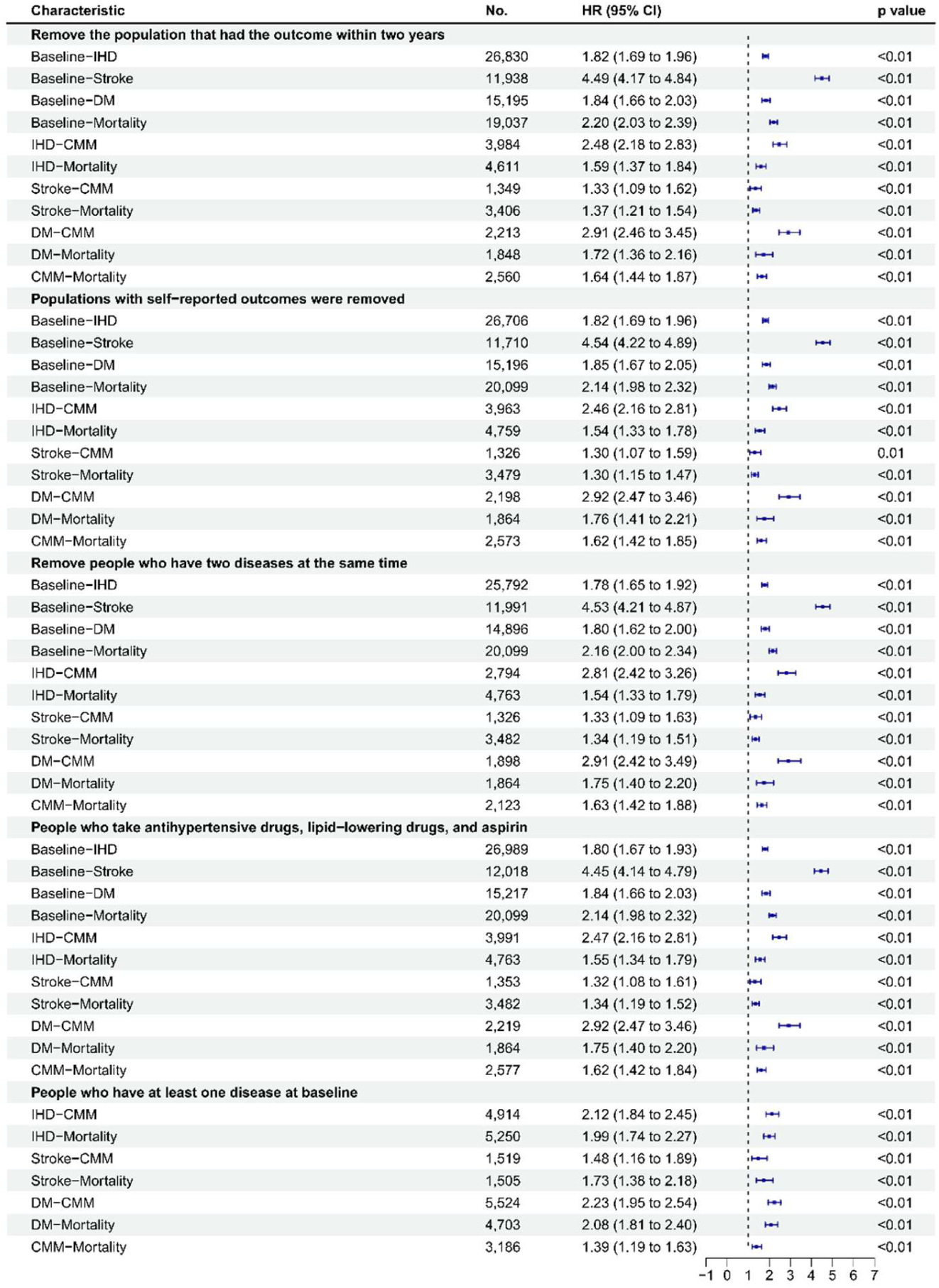
Sensitivity analyses included (1) remove the population that had the outcome within two years (2) populations with self-reported outcomes were removed; (3) remove people who have two diseases at the same time (4) including people who take antihypertensive drugs, lipid-lowering drugs and aspirin; (5)including participants with a prior diagnosis of heart disease, stroke, or diabetes were included and assigned to CMD or CMM status based on their CMD status at baseline.HR, hazard ratio; CI, confidence interval; IHD, ischemic heart disease; DM, diabetes mellitus; CMM, cardiometabolic multimorbidity.

**Figure 4.**
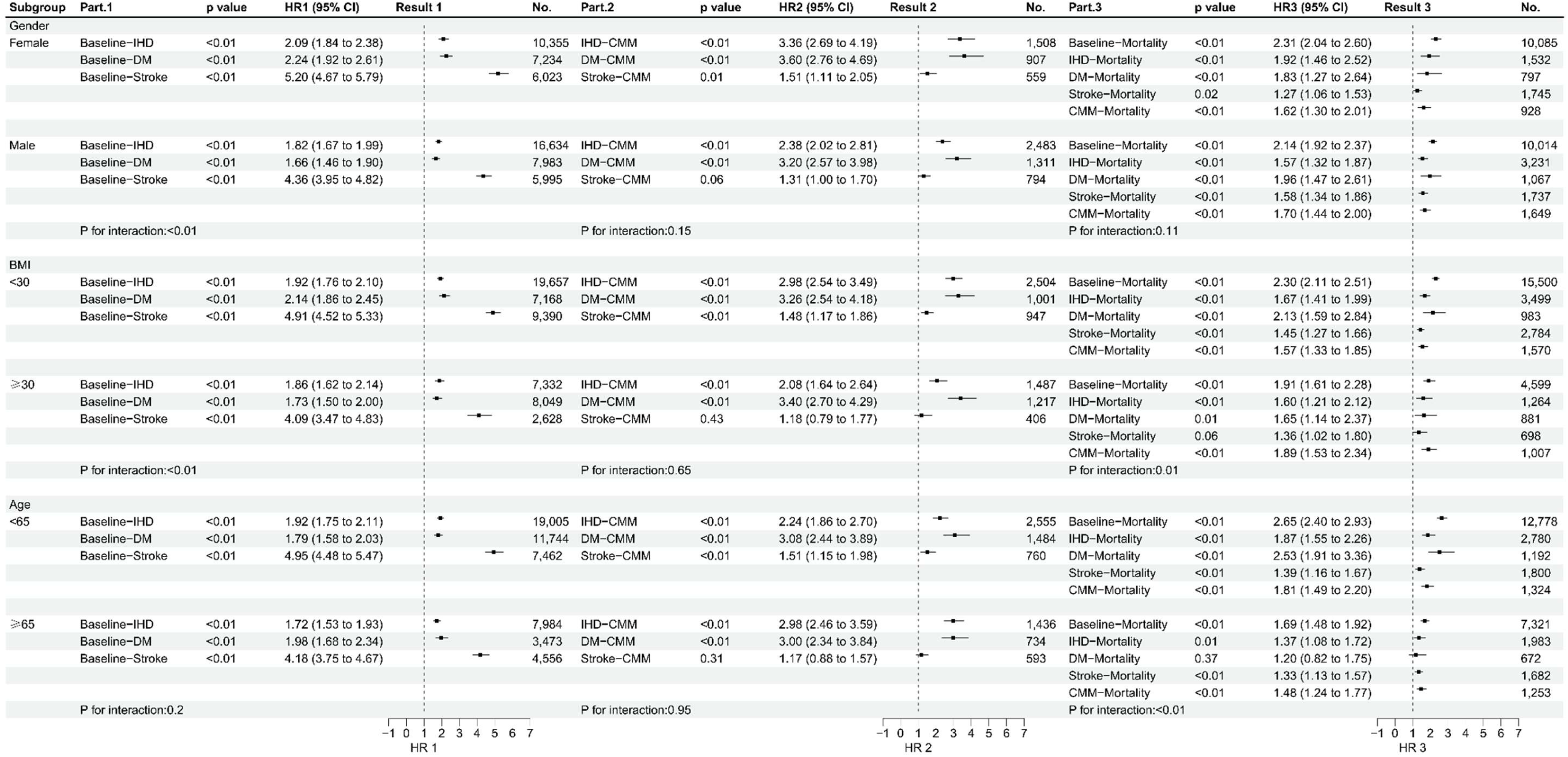
Subgroup analyses were conducted across all transition pathways, stratified by age (e.g., <65 vs. ≥65 years), sex (male vs. female), and BMI categories (normal vs. obese). HR, hazard ratio; CI, confidence interval; CMM, cardiometabolic multimorbidity; IHD, ischemic heart disease; DM, diabetes mellitus.

## Discussion

This study included 366,616 eligible participants and demonstrated that TBI exerted significant adverse effects across all stages of disease progression from health to CMM and mortality. In multi-state models, we further identified that TBI exhibited differential impacts on individual CMD components at each transition stage. These findings remained robust in most subgroup analyses and after adjustment for multiple potential confounders.

In line with prior studies that demonstrated an increased risk of isolated CMD in participants with TBI, our results further confirmed that individuals with TBI were associated with 72%, 90%, and 120% increased risks of IHD, DM, and stroke, respectively, compared with those without TBI.^12, 22–25^ These estimates were partially aligned with findings from our Cox proportional hazards models and multi-state models. Furthermore, we examined the associations of TBI with CMM and mortality, and determined that TBI was associated with 3.98-fold higher risk of CMM and 2.57-fold higher risk of all-cause mortality relative to TBI-free participants. Meanwhile, we observed that the strength of association between TBI and the outcomes of CMM, stroke, and mortality was substantially stronger than that for IHD and DM. Accordingly, early stroke screening and monitoring among patients with TBI may hold greater clinical value for preventing progression to CMM and reducing the risk of all-cause mortality. Furthermore, previous studies have explored potential mechanisms underlying the associations between TBI and CMD/CMM. These include TBI-related cardiovascular autonomic dysfunction,^26, 27^ insulin resistance,^28^ and elevated stroke risk mediated by alterations in neuroimmune and neurometabolic functions. ^29, 30^ In recent years, an alternative mechanistic hypothesis has also been proposed: the association of TBI with multiple CMDs may be driven by brain–gut axis dysfunction induced by TBI-triggered neuroinflammation and neurodegeneration.^31–33^ These findings suggest that the effects of TBI on CMD and CMM are likely multifactorial. Prior research has conceptualized TBI as a chronic condition and identified potential bidirectional interactions between TBI and CMD.^34, 35^ For instance, one study reported that patients with TBI had a 1.35–fold higher risk of myocardial dysfunction than individuals without TBI,^36^ with synergistic interplay involving neuroinflammation and vascular dysregulation also hypothesized.^17^ Nevertheless, few studies have systematically examined differential effects of TBI on the progression of distinct CMD subtypes to CMM. Accordingly, in contrast to previous investigations, the present study employed multi-state models to further examine the role of TBI across the entire disease continuum, spanning from a healthy state to incident CMD, subsequent progression to CMM, and eventual mortality. This analytical framework not only allowed direct comparison of TBI-associated risk estimates across different CMD subtypes but also enabled detailed assessment of how TBI influences transitions from individual CMDs to CMM or mortality. During the transition from health to incident CMD, the hazard ratio for TBI in relation to stroke was substantially higher than that for IHD and DM, a pattern consistent with findings from Cox proportional hazards models. However, during the transition from CMD to CMM, although TBI remained statistically significantly associated with an elevated risk of progression to CMM among individuals with IHD, DM, or stroke, the hazard ratios of TBI for incident CMM in those with pre-existing IHD and DM were higher than the corresponding hazard ratios for progression from baseline health to the respective CMD events. By contrast, the pattern of hazard ratios linking TBI to stroke onset and subsequent progression from stroke to CMM was completely reversed. Integrating these findings with the aforementioned mechanistic evidence and the main results of this study, we hypothesize that TBI confers a particularly high baseline risk of stroke. Therefore, among participants who initially developed other forms of CMD, TBI may further increase stroke risk through overlapping pathophysiological mechanisms and shared risk factors, which in turn raises the hazard ratio for progression to CMM. Meanwhile, for participants who have already experienced stroke, TBI and stroke may exert similar systemic pathophysiological effects. As such, a subsequent TBI event may not substantially alter lifestyle or long-term prognosis in patients with pre-existing stroke. Consequently, during the progression from CMD to CMM, the hazard ratio of TBI for transition to CMM among stroke survivors is notably less prominent than that observed among individuals with other CMD subtypes. This study is therefore the first to systematically explore the predictive value of TBI for subsequent progression to CMM or all-cause mortality among patients with DM, stroke, or IHD within the same population-based cohort. Multiple sensitivity analyses further confirmed the robustness of the observed associations between TBI and the sequential transition stages from health to distinct CMDs, then to CMM and mortality. Collectively, these findings highlight that targeted and effective interventions for patients with TBI—regardless of whether they have pre-existing CMD—carry substantial clinical value for preventing progression to CMM and reducing all-cause mortality. Finally, in subgroup analyses stratified by BMI and age, the impact of TBI on multiple transition stages—from a healthy state to CMD or all-cause mortality, and from CMD to mortality—was weaker among individuals with BMI ≥ 30 kg/m² and aged ≥ 65 years than among their counterparts with BMI < 30 kg/m² and aged < 65 years. Integrating this observation with previous TBI-related research^7, 8^ and baseline profiles of physical activity and dietary habits in the current study, we hypothesize that TBI may reduce patients’ exercise capacity and disrupt their lifestyle. Since individuals with BMI ≥ 30 kg/m² and aged ≥ 65 years already possess poorer baseline exercise capacity than younger and non-obese individuals, the additional detrimental effect of TBI appears less pronounced in this subgroup. This further implies that one plausible pathway through which TBI affects the sequential transitions from health to distinct CMDs, then to CMM and mortality, may involve TBI-induced changes in exercise capacity and lifestyle. Accordingly, early rehabilitation support and prompt restoration of functional ability following TBI may lower the risks of subsequent CMD, CMM, and all-cause mortality among patients with TBI.

Key strengths of this study include the direct comparison of TBI-related risk estimates across multiple CMD subtypes within a single cohort, which provides critical evidence for developing targeted preventive strategies for patients with distinct CMD phenotypes. Additional strengths include its large sample size and rigorous prospective cohort design, which enhance the reliability and generalizability of the findings. Nevertheless, this study has several limitations. First, a proportion of participants were excluded due to missing baseline data or other eligibility constraints. While we adjusted for as many potential confounders as possible in multi-state models, residual confounding may still affect the precision of risk estimate comparisons. Second, the UKB database is subject to healthy participant bias, which restricts this study to identifying correlational associations between TBI and the study outcomes, rather than establishing definitive causal relationships. Third, CMD ascertainment based on ICD-10 coding is susceptible to administrative variability. Although we excluded self-reported diagnoses in sensitivity analyses to reduce misclassification bias, minor measurement bias in risk comparisons cannot be fully eliminated.

In conclusion, this large prospective cohort study demonstrates that TBI exerts differential effects on disease progression from a healthy state to CMD, as well as subsequent transitions to CMM and mortality, with distinct impacts on specific CMD transition pathways. Further research is warranted to elucidate the underlying biological mechanisms and establish causal relationships between TBI and cardiometabolic deterioration. Collectively, our findings confirm TBI as a critical risk factor for CMD, CMM, and all-cause mortality, highlighting that early intervention and targeted management of TBI patients may improve long-term clinical prognoses.

## Data Availability

All the data used in this study were derived from UK Biobank (https://www.ukbiobank.ac.uk/). This study was conducted using the UK Biobank Resource under Application 84443.

## Acknowledgements

Not applicable

## Disclosures

### Ethics approval and consent to participate

The study was approved by the North West Multi-Centre Research Ethics Committee, conducted in accordance with the Helsinki Declaration of 1975, as revised in 2000. Informed consent was obtained from all individual participants included in the study.

### Consent for publication

Not applicable

### Authors’ contributions

S.L., X.B.L, X.J.C., W.X. contributed to the study concept and design; acquisition, analysis, and interpretation of the data; drafting and critical revision of the manuscript. X.Y.C., Y.R.L., S.Y.L, L.L. contributed to the study concept and design; acquisition, analysis. Y.P.B., C.C.L. contributed to the acquisition and interpretation of the data and critical revision of the manuscript. All authors gave final approval of the version to be published. X.J.C. is the guarantor of this work and, as such, had full access to all the data in the study and takes responsibility for the integrity of the data and accuracy of data analysis.

## Abbreviations

TBI: traumatic brain injury
CMD: cardiometabolic disease
CMM: cardiometabolic multimorbidity
UKB: UK Biobank
DM: diabetes mellitus
IHD: ischemic heart disease
CVD: cardiovascular disease
EHRs: electronic health records
LDL: low density lipoprotein
CHOL: Cholesterol
BMI: body mass index
SBP: systolic blood pressure
DBP: diastolic blood pressure

---

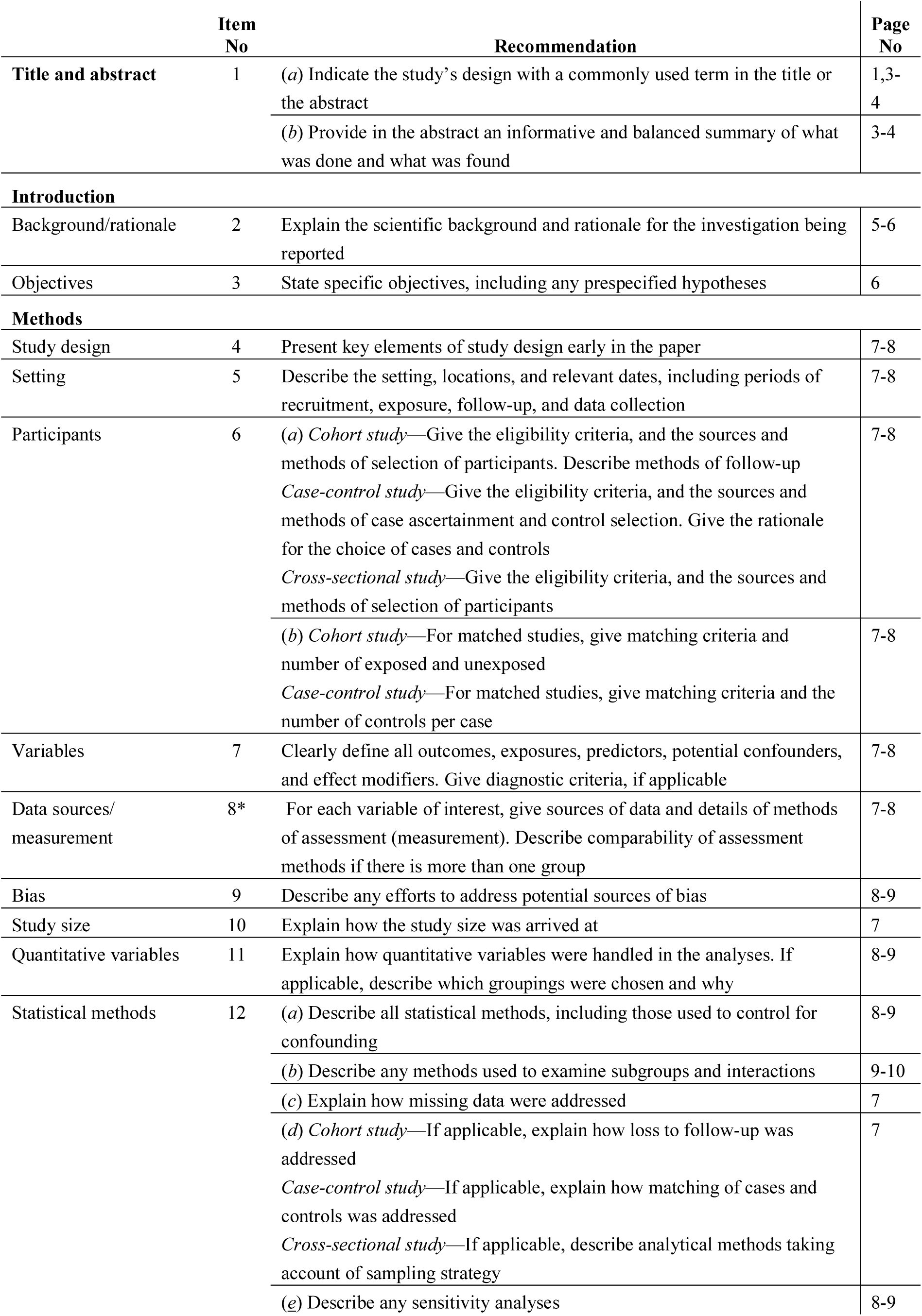
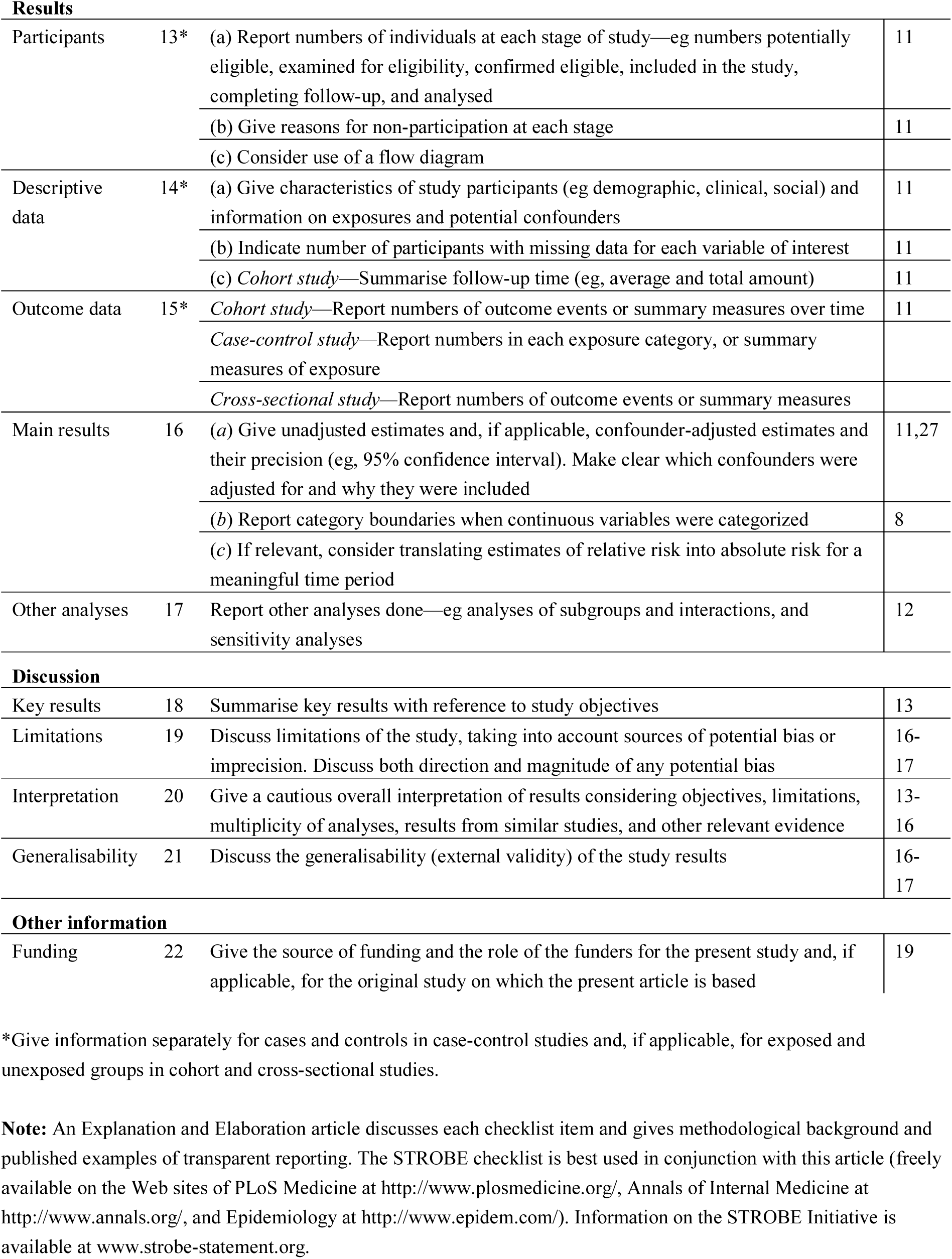
STROBE Statement—checklist of items that should be included in reports of observational studies.

|  | Item No | Recommendation | Page No |
| --- | --- | --- | --- |
| Title and abstract | 1 | (a) Indicate the study’s design with a commonly used term in the title or the abstract | 1,3-4 |
|  |  | (b) Provide in the abstract an informative and balanced summary of what was done and what was found | 3-4 |
| Introduction |  |  |  |
| Background/rationale | 2 | Explain the scientific background and rationale for the investigation being reported | 5-6 |
| Objectives | 3 | State specific objectives, including any prespecified hypotheses | 6 |
| Methods |  |  |  |
| Study design | 4 | Present key elements of study design early in the paper | 7-8 |
| Setting | 5 | Describe the setting, locations, and relevant dates, including periods of recruitment, exposure, follow-up, and data collection | 7-8 |
| Participants | 6 | (a) Cohort study—Give the eligibility criteria, and the sources and methods of selection of participants. Describe methods of follow-up<br>Case-control study—Give the eligibility criteria, and the sources and methods of case ascertainment and control selection. Give the rationale for the choice of cases and controls<br>Cross-sectional study—Give the eligibility criteria, and the sources and methods of selection of participants | 7-8 |
|  |  | (b) Cohort study—For matched studies, give matching criteria and number of exposed and unexposed<br>Case-control study—For matched studies, give matching criteria and the number of controls per case | 7-8 |
| Variables | 7 | Clearly define all outcomes, exposures, predictors, potential confounders, and effect modifiers. Give diagnostic criteria, if applicable | 7-8 |
| Data sources/<br>measurement | 8* | For each variable of interest, give sources of data and details of methods of assessment (measurement). Describe comparability of assessment methods if there is more than one group | 7-8 |
| Bias | 9 | Describe any efforts to address potential sources of bias | 8-9 |
| Study size | 10 | Explain how the study size was arrived at | 7 |
| Quantitative variables | 11 | Explain how quantitative variables were handled in the analyses. If applicable, describe which groupings were chosen and why | 8-9 |
| Statistical methods | 12 | (a) Describe all statistical methods, including those used to control for confounding | 8-9 |
|  |  | (b) Describe any methods used to examine subgroups and interactions | 9-10 |
|  |  | (c) Explain how missing data were addressed | 7 |
|  |  | (d) Cohort study—If applicable, explain how loss to follow-up was addressed<br>Case-control study—If applicable, explain how matching of cases and controls was addressed<br>Cross-sectional study—If applicable, describe analytical methods taking account of sampling strategy | 7 |
|  |  | (e) Describe any sensitivity analyses | 8-9 |
| **Results** |  |  |  |
| Participants | 13* | (a) Report numbers of individuals at each stage of study—eg numbers potentially eligible, examined for eligibility, confirmed eligible, included in the study, completing follow-up, and analysed | 11 |
|  |  | (b) Give reasons for non-participation at each stage | 11 |
|  |  | (c) Consider use of a flow diagram |  |
| Descriptive data | 14* | (a) Give characteristics of study participants (eg demographic, clinical, social) and information on exposures and potential confounders | 11 |
|  |  | (b) Indicate number of participants with missing data for each variable of interest | 11 |
|  |  | (c) *Cohort study*—Summarise follow-up time (eg, average and total amount) | 11 |
| Outcome data | 15* | *Cohort study*—Report numbers of outcome events or summary measures over time | 11 |
|  |  | *Case-control study*—Report numbers in each exposure category, or summary measures of exposure |  |
|  |  | *Cross-sectional study*—Report numbers of outcome events or summary measures |  |
| Main results | 16 | (a) Give unadjusted estimates and, if applicable, confounder-adjusted estimates and their precision (eg, 95% confidence interval). Make clear which confounders were adjusted for and why they were included | 11,27 |
|  |  | (b) Report category boundaries when continuous variables were categorized | 8 |
|  |  | (c) If relevant, consider translating estimates of relative risk into absolute risk for a meaningful time period |  |
| Other analyses | 17 | Report other analyses done—eg analyses of subgroups and interactions, and sensitivity analyses | 12 |
| **Discussion** |  |  |  |
| Key results | 18 | Summarise key results with reference to study objectives | 13 |
| Limitations | 19 | Discuss limitations of the study, taking into account sources of potential bias or imprecision. Discuss both direction and magnitude of any potential bias | 16-17 |
| Interpretation | 20 | Give a cautious overall interpretation of results considering objectives, limitations, multiplicity of analyses, results from similar studies, and other relevant evidence | 13-16 |
| Generalisability | 21 | Discuss the generalisability (external validity) of the study results | 16-17 |
| **Other information** |  |  |  |
| Funding | 22 | Give the source of funding and the role of the funders for the present study and, if applicable, for the original study on which the present article is based | 19 |

\*Give information separately for cases and controls in case-control studies and, if applicable, for exposed and unexposed groups in cohort and cross-sectional studies.
**Note:** An Explanation and Elaboration article discusses each checklist item and gives methodological background and published examples of transparent reporting. The STROBE checklist is best used in conjunction with this article (freely available on the Web sites of PLoS Medicine at http://www.plosmedicine.org/, Annals of Internal Medicine at http://www.annals.org/, and Epidemiology at http://www.epidem.com/). Information on the STROBE Initiative is available at www.strobe-statement.org.

